# Potential use of antibodies to provide an earlier indication of lymphatic filariasis resurgence in post-mass drug administration surveillance, American Samoa

**DOI:** 10.1101/2021.11.29.21267031

**Authors:** Angela M. Cadavid Restrepo, Katherine Gass, Kimberly Y. Won, Meru Sheel, Keri Robinson, Patricia M. Graves, Saipale Fuimaono, Colleen L Lau

## Abstract

**Objectives:** Under the Global Programme to Eliminate Lymphatic Filariasis (LF), American Samoa conducted seven rounds of mass drug administration between 2000 and 2006. The territory passed transmission assessment surveys (TAS) in 2011 (TAS-1) and 2015 (TAS-2) based on World Health Organization guidelines. In 2016, the territory failed TAS-3, indicating resurgence. This study aims to determine if antibodies (Ab) may have provided a timelier indication of LF resurgence in American Samoa.

**Methods:** We examined school-level Ag and Ab status (presence/absence of Ag- and Ab-positive children) and prevalence of single and combined Ab responses to Wb123, Bm14, Bm33 Ags at each TAS. Pearson’s chi-squared tests and logistic regression were used to examine associations between school-level Ab prevalence in TAS-1 and TAS-2 and school-level Ag status in TAS-3.

**Results:** Schools with higher prevalence of Wb123 Ab in TAS-2 had higher odds of being Ag-positive in TAS-3 (odds ratio [OR] 24.5, 95% CI:1.2-512.7). Schools that were Ab-positive for WB123 plus Bm14, Bm33 or both Bm14 and Bm33 in TAS-2 had higher odds of being Ag-positive in TAS-3 (OR 16.0-24.5).

**Conclusion:** Anti-filarial Abs could provide earlier signals of resurgence and enable a timelier response. The promising role of Abs in post-MDA surveillance and decision making should be further investigated in other settings.

## Introduction

Lymphatic filariasis (LF) is a parasitic infection caused by three species of the filarial nematodes, *Wuchereria bancrofti, Brugia malayi* and *Brugia timori*, that are transmitted between definitive human hosts by multiple mosquito vectors (*Culex, Anopheles, Aedes*, and *Mansonia*) (Centers for Disease Control and Prevention (CDC)). In 2000, in response to the World Health Assembly resolution WHA50.29, the World Health Organization (WHO) targeted LF for global elimination by 2020 and launched the Global Programme to Eliminate Lymphatic Filariasis (GPELF) (World Health Organization, 2000). One of the strategies proposed by GPELF focused on interrupting transmission by implementing mass drug administration (MDA) of antifilarial drugs in endemic areas (World Health Organization, 2000). A key challenge faced by most LF-endemic countries that have implemented MDA is to effectively undertake post-validation surveillance (Lau et al., 2020).

Transmission Assessment Surveys (TAS) are recommended by WHO in children aged 6-7 years in geographically defined evaluation units as the tool to measure the impact of MDA and determine whether the targets have been reached (World Health Organization, 2011). TAS are school based if attendance is high; otherwise done in community cluster surveys. MDA is stopped when infection prevalence has been reduced to a level where it is presumed that transmission cannot be sustained even in the absence of further interventions. Estimates suggest that 4-6 annual rounds of MDA with effective population coverage (>65% of the total population) are required to reduce antigen (Ag) prevalence to below 2% in areas where *Anopheles* or *Culex* is the main vector, and 1% where *Aedes* is the dominant vector (World Health Organization, 2011). TAS is a population-based survey designed to estimate the prevalence of Ag (in *W. bancrofti*-endemic areas) or antifilarial antibodies (Ab) (in Brugian endemic areas) in children aged 6 to 7 years. This age group was selected because new incident infections would reflect recent exposure to ongoing transmission (World Health Organization, 2011). According to WHO guidelines, TAS should also be repeated at 2-3 and 4-6 years after stopping MDA in each evaluation unit to monitor and identify signals of resurgence (World Health Organization, 2011).

In American Samoa an MDA programme to eliminate LF was initiated in 2000 under the Pacific Programme for the Elimination of Lymphatic Filariasis (World Health Organization, 2006). Seven rounds of MDA with a single dose of diethylcarbamazine (DEC) and albendazole were conducted between 2000 and 2006 (World Health Organization, 2006). In the first three years, population coverage by MDA was 24-52% and improved to 65-71% in the subsequent four years (World Health Organization, 2006). American Samoa passed TAS-1 (in February 2011) and TAS-2 (in April 2015) with numbers of Ag-positive children below the critical cut-off of six (two Ag-positive children in TAS-1 and one Ag-positive child in TAS-2, equivalent to crude prevalence of 0.2% (95% CI 0.0 to 0.8%), and 0.1% (95% CI 0.0 to 0.7%), respectively (Won Kimberly Y. et al., 2018). However, the territory failed TAS-3 in November 2016, with nine Ag-positive children, an adjusted prevalence of 0.7% (95% CI 0.3 to 1.8%), higher than the cut-off and the recommended upper confidence limit of 1% (Sheel et al., 2018). In addition, evidence from other human and entomological research surveys conducted in 2010 and 2014 suggested ongoing LF transmission and the persistence of residual hotspots (Lau et al., 2017, Lau et al., 2014, Lau et al., 2016, Schmaedick et al., 2014). In 2016, in parallel with TAS-3, a community survey of residents aged ≥8 years confirmed LF resurgence with adjusted Ag prevalence of 6.2% (95% CI: 4.5-8.6%). Spatial analyses of the 2016 community survey data also identified the potential existence of new or previously unidentified LF hotspots in the territory (Lau et al., 2020).

There is a current need to strengthen post-MDA surveillance through the development of alternative or additional surveillance strategies to identify residual LF infections and ensure long-term success of MDA. As LF elimination programs progress towards the end stages, one of the key challenges is the availability of diagnostics that are sufficiently sensitive for detecting low level transmission or resurgence. Ag prevalence declines after the implementation of MDA, and as prevalence drops to low levels, more accurate tests and surveillance methods will be required to detect transmission signals. TAS currently relies on Ag test results only, and antigenemia alone may not be sensitive enough to ensure timely detection of ongoing transmission or recrudescence (Lau et al., 2020). Existing evidence suggests that Ab responses may provide an earlier indicator of filarial infection than Ag (Joseph H et al., 2011). While the development and duration of serological responses to specific anti-filarial Abs such as Bm14, Bm33 and Wb123 are poorly understood, Ab testing may have a potential role in post-MDA and post-validation surveillance in *W. bancrofti*-endemic areas (Won Kimberly Y et al., 2018).

Here, we examined the potential to use combinations of Ag and Ab tests as surveillance markers to provide earlier signals of transmission. This study aimed to geographically visualise and compare LF Ag and Ab signals in American Samoa at the school level for TAS-1, TAS-2, and TAS-3, and to determine if anti-filarial Abs in TAS-1 and TAS-2 may have provided an earlier indication of areas at risk for ongoing transmission in American Samoa.

## Methods

### A. Ethics statement

Ethics approvals for TAS-1 and TAS-2 were granted by the relevant committees and are reported in previously published work (Won Kimberly Y. et al., 2018). TAS-3 was approved by the American Samoa Institutional Review Board and the Human Research Ethics Committee at the Australian National University (protocol number 2016/482). Full details of collaborations and official permissions for school and village visits in 2016 have been previously described (Sheel et al., 2018).

### B. Study setting

American Samoa is a United States territory in the South Pacific comprised of five inhabited islands: Tutuila, Aunu’u, Ofu, Olosega and Ta’ū, and two uninhabited coral atolls (Figure 1). The population was 55,519 in 2010, the majority of whom lived in the largest island, Tutuila (United States Census Bureau, 2010). The population is young, with one-third aged under 15 years (United States Census Bureau, 2010). Education is compulsory between ages 6 and 18 and is provided by public and private elementary and secondary schools (Amerika Samoa Department of Education, 2020).

**Figure 1.**
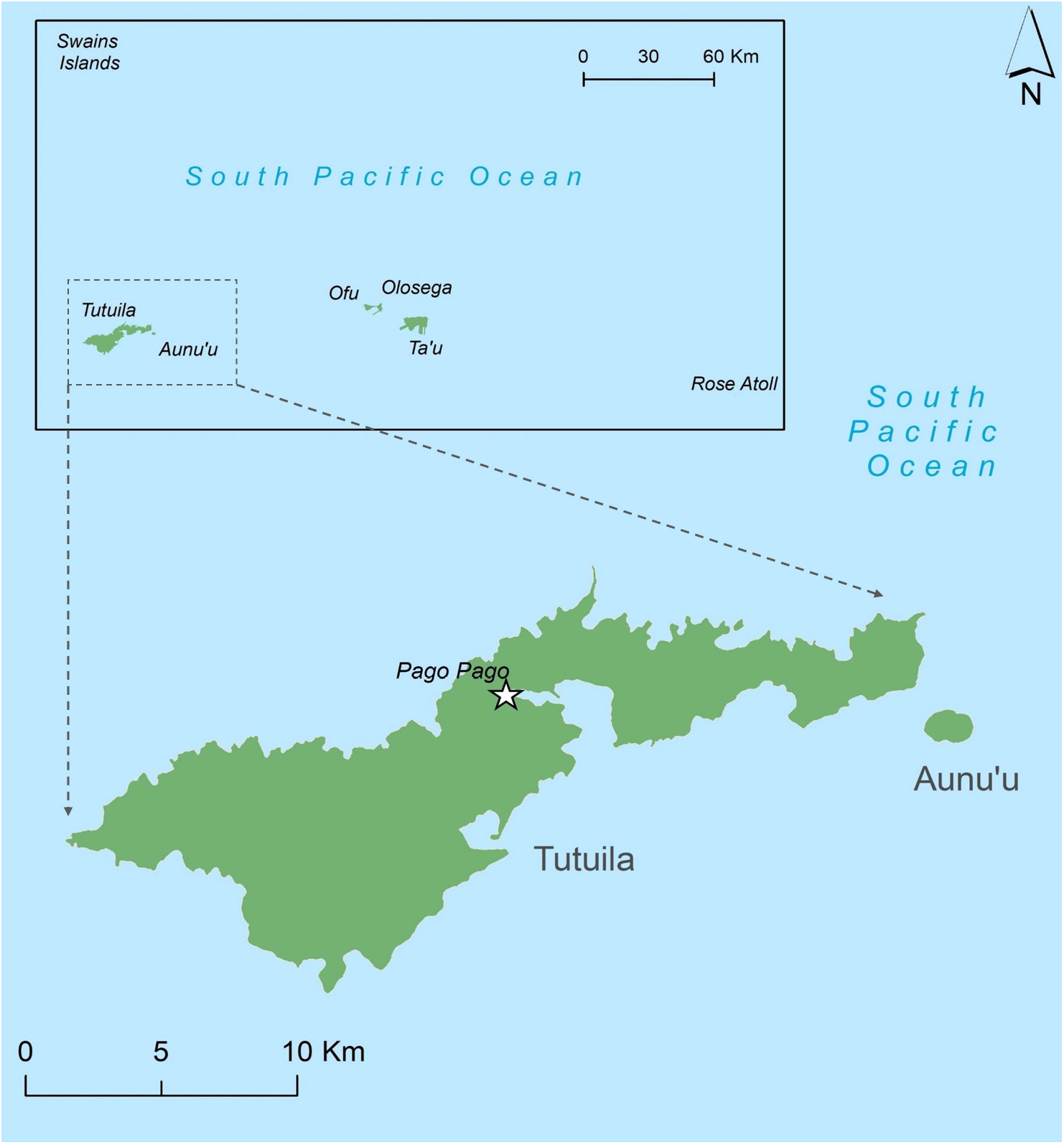
Map of American Samoa

In American Samoa, LF is caused by *W. bancrofti* which are diurnally sub-periodic worms transmitted predominantly by the highly efficient day-biting mosquito *Aedes polynesiensis*, and also night-biting *Aedes samoanus* as a secondary vector (Schmaedick et al., 2014).

### C. Data sources

Data were obtained from three TAS conducted across American Samoa in 2011, 2015 and 2016. Surveys were carried out at 25, 30 and 29 schools for each survey year, respectively (all elementary schools on the main island of Tutuila and the adjacent island of Aunu’u). Since each TAS included children who attended Grades 1 and 2 (used as a proxy for being 6-7 years old), each survey was conducted in a different cohort of children.

Informed written consent was obtained from a parent or guardian. Finger prick blood samples (200µL) were used to detect circulating filarial Ag. Binax NOW® Filariasis Immunochromatographic test (ICT) (Alere, Scarborough, ME) was used in TAS-1 and TAS-2, and Alere^™^ Filariasis Test Strip (FTS) (Abbott, Scarborough, ME) in TAS-3 (Abbott, Weil et al., 2013). Dried blood spots were prepared for later elution and antibody testing, where anti-filarial Ab responses were tested by luciferase immunoprecipitation system (LIPS) assay (for IgG responses to Wb123 in TAS-1) or multiplex bead assay (MBA) (for anti-filarial responses to Bm14 and Bm33 in TAS-1, and Wb123, Bm14 and Bm33 in TAS-2 and TAS-3) (Kubofcik et al., 2012, Lammie et al., 2012). A minimum of four controls were used for internal quality control for these TAS MBA analyses. The first was a buffer blank that contained only the assay buffer, which was used to subtract out any background noise. At least two controls were pools of reference sera that served as known positives for Abs to be detected in the assay. The number of positive controls included varied due to total number of Ag included on the MBA. The concentration of referent sera added was adjusted so one control represented high positives near the maximum value the instrument can detect and the other control represented low positives in the linear range of detection. The last control was a negative control with known negative LF-status. For TAS-1 and-2, the cut-off determination methods have been previously described (Won Kimberly Y. et al., 2018). Receiver Operating Characteristic (ROC) curves were calculated using sera from *W. bancrofti*-infected patients and presumed negative sera from US citizens with no history of foreign travel. For TAS-3 cut-off determination, the mean plus three standard deviations (SDs) method was used, as it has been previously described (Moss et al., 2011, Priest et al., 2016). Eighty-six serum samples collected from healthy North American adults (where LF is not endemic) with no history of foreign travel were run using all the same conditions and reagents as the study samples. A mean plus three SDs was calculated and used as a cut-off whereby any mean fluorescent intensity value at or above was considered positive and below considered negative.

Parents/guardians of children found to be Ag-positive were informed, and participants were offered a standard single dose of DEC (6 mg/kg) and albendazole (400 mg). Full details of survey designs and data collection have been reported elsewhere (Sheel et al., 2018, Won Kimberly Y. et al., 2018).

An administrative boundary map was downloaded from the American Samoa Coastal and Marine Spatial Planning Data Portal (Marine Cadastre Admin, 2020). During TAS-3, the geographical coordinates of each school were collected using a hand-held global positioning system (Sheel et al., 2018). and imported into ArcGIS version 10.7.1 to create a shapefile of all elementary schools (ESRI: Environmental Systems Research Institute, 2019).

### D. Data analysis

A total of 33 schools were included in at least one of the surveys. Some elementary schools that participated in TAS-1, TAS-2, or in both, were closed and new schools were opened in the same/similar geographic area by the time TAS-3 was conducted. Thus, Vatia and Mt. Alava Elementary Schools located in the small village of Vatia; Olomoana and Aoa schools located in Aoa village; and Iakina Adventist Academy and SDA located in Ili’ili village were considered as the same school in the analyses.

Because Ag and Ab prevalence at school level was low, the school Ag and Ab status was used for most analyses. Ag-positive schools were defined as those with at least one Ag-positive child. Ab-positive schools were defined as those with at least one child who tested positive to a single Ab or different combinations of Ab responses. For each TAS, summary statistics were calculated for the whole survey and for the 30 school locations. School-level crude prevalence of Ag, single Abs and different combinations of Wb123, Bm14 and Bm33 Abs (see below) were estimated, and binomial exact methods were applied to estimate 95% confidence intervals (95% CI). Bar plots were created to show the prevalence of Ab-positive children in TAS-1 and TAS-2 stratified by the school Ag status (Ag-positive or Ag-negative) in TAS-3.

To enable comparisons over time, five schools that were not included in all three TAS were excluded from direct comparisons (Le’atele (Fagasa), Pacific Horizon, Peteli Academy, St. Theresa and Ta’iala Academy). Therefore, only 25 school locations were included in the final analyses. The school Ab status in TAS-1 and TAS-2 and the school Ag status in TAS-3 were compared using Pearson’s chi squared tests. The following combinations of Ab responses at the school level in TAS-1 and TAS-2 were also examined to assess the value of testing a combination of Abs on diagnostic performance: i) positive response to at least one Ab in the combinations, denoted from here as Wb123∪Bm14, Wb123∪Bm33, Bm14∪Bm33 and Wb123∪Bm14∪Bm33, and ii) positive response to all Abs in the combinations, denoted from here as Wb123∩Bm14, Wb123∩Bm33, Bm14∩Bm33 and Wb123∩Bm14∩Bm33. Sensitivity, specificity, positive predictive value (PPV) and negative predictive value (NPV) of Ab-positive schools in TAS-1 and TAS-2 for predicting Ag-positive schools in TAS-3 were also estimated.

Univariate logistic regression analyses were conducted to examine associations between single and combinations of Ab responses in TAS-1 and TAS-2 at the school level and school Ag status in TAS-3. Haldane’s correction for odds ratio was used when either all children were Ag-positive or all were Ag-negative for a particular Ab response or combination of Ab responses (a value of 0.5 was added to every cell when cross□product ratios of a 2 × 2 contingency table was zero) (Haldane, 1940, Lawson, 2004).

In all analyses, statistical significance was determined with α-levels of 0.05 (as indicated by 95% CI). All analyses were conducted using R software version R-4.0.3 (R Core Team, 2020). Data were imported into ArcGIS version 10.7.1 (ESRI: Environmental Systems Research Institute, 2019) and linked spatially to the surveyed schools to generate maps that show the geographical distribution of the surveyed schools, the number of Ag-positive children identified through TAS, and crude Ab prevalence for each school.

## Results

The initial data set consisted of 33 schools and a total of 1,134 elementary school children that participated in TAS-1, 864 in TAS-2 and 1,143 in TAS-3. A summary of TAS participants and results is presented in Table 1. The overall crude Ag prevalence were 0.2% (95% CI: 0-0.8%) in TAS-1 (n=937), 0.1% (95% CI: 0-0.7%) in TAS-2 (n=768), and 0.8% (95% CI: 0.4-1.5%) in TAS-3 (n=1,143). After adjusting for survey design, age and sex distribution, Ag prevalence was 0.7% (95% CI: 0.3-1.8%) in TAS-3 (Sheel et al., 2018). Supplementary table 1 shows crude Ag prevalence for each school in the three surveys. Lupelele Elementary was the only school with Ag-positive school children in TAS-1 (two positive children out of 92 tested, Ag prevalence 2.2%, 95% CI: 0.26%-7.63%) and TAS-2 (one positive child out of 85 tested, Ag prevalence 1.2%, 95% CI: 0%-6.4%). Also, Lupelele Elementary was among five schools with Ag-positive children in TAS-3. In TAS-3, Ag prevalence by school ranged from 0%-4.9%; Coleman Elementary had the highest number of Ag-positive students (Supplementary table 1). School locations included in the three surveys and the number of Ag-positive children identified through TAS are shown in Figure 2.

**Table 1.** Summary of participants and results from TAS-1 (2011), TAS-2 (2015) and TAS-3 (2016) surveys in American Samoa.

|  | **TAS-1** | **TAS-2** | **TAS-3** |
| --- | --- | --- | --- |
| **Timing** | February 2011 | April 2015 | September 2016 |
| **Total number of schools** | 25 | 30 | 29 |
| **Total number of participants** | 1,134 | 864 | 1,143 |
| **Number of participants with valid Ag test results** | 937 | 768 | 1,143 |
| **Number of Ag-positive children** | 2 | 1 | 9 |
| **Crude Ag prevalence (95% CI)** | 0.2% (0.0-0.8) | 0.1% (0.0-0.7) | 0.7% (0.4-1.5) |
| **Number of participants with Ab test results** | 1,112 | 836 | 1,139 |
| **Number of Ab-positive children and crude prevalence (%)** |  |  |  |
| **Wb123** | 11(1.0%) | 30 (3.6%) | 94 (8.3%) |
| **Bm14** | 76 (6.8%) | 25 (3.0%) | 18 (1.6%) |
| **Bm33** | 133 (12.0%) | 65 (7.8%) | 237 (20.8%) |
| **Number and % of schools with at least one Ab-positive child** |  |  |  |
| **Wb123** | 6 (24.0%) | 12 (40.0%) | 22 (75.9%) |
| **Bm14** | 22 (88.0%) | 12 (40.0%) | 10 (34.5%) |
| **Bm33** | 20 (80.0%) | 17 (56.7%) | 25 (86.2%) |
| **Number and % of schools that were positive for at least one Ab in the following combinations** |  |  |  |
| **Wb123∪Bm14** | 22 (88.0%) | 14 (46.6%) | 21 (72.4%) |
| **Wb123∪Bm33** | 20 (80.0%) | 17 (56.6%) | 24 (82.8%) |
| **Bm14∪Bm33** | 25 (100.0%) | 18 (60.0%) | 22 (75.9%) |
| **Wb123∪Bm14∪Bm33** | 25 (100 %) | 18 (60.0%) | 24 (82.6%) |
| **Number and % of schools that were positive for all Abs in the following combinations** |  |  |  |
| **Wb123∩Bm14** | 6 (24.0%) | 8 (26.7%) | 9 (31.0%) |
| **Wb123∩Bm33** | 6 (24.0%) | 11 (36.7%) | 20 (67.0%) |
| **Bm14∩Bm33** | 17 (68.0%) | 10 (33.3%) | 10 (34.5%) |
| **Wb123∩Bm14∩Bm33** | 6 (24.0%) | 8 (26.7%) | 9 (31.0%) |

**Figure 2.**
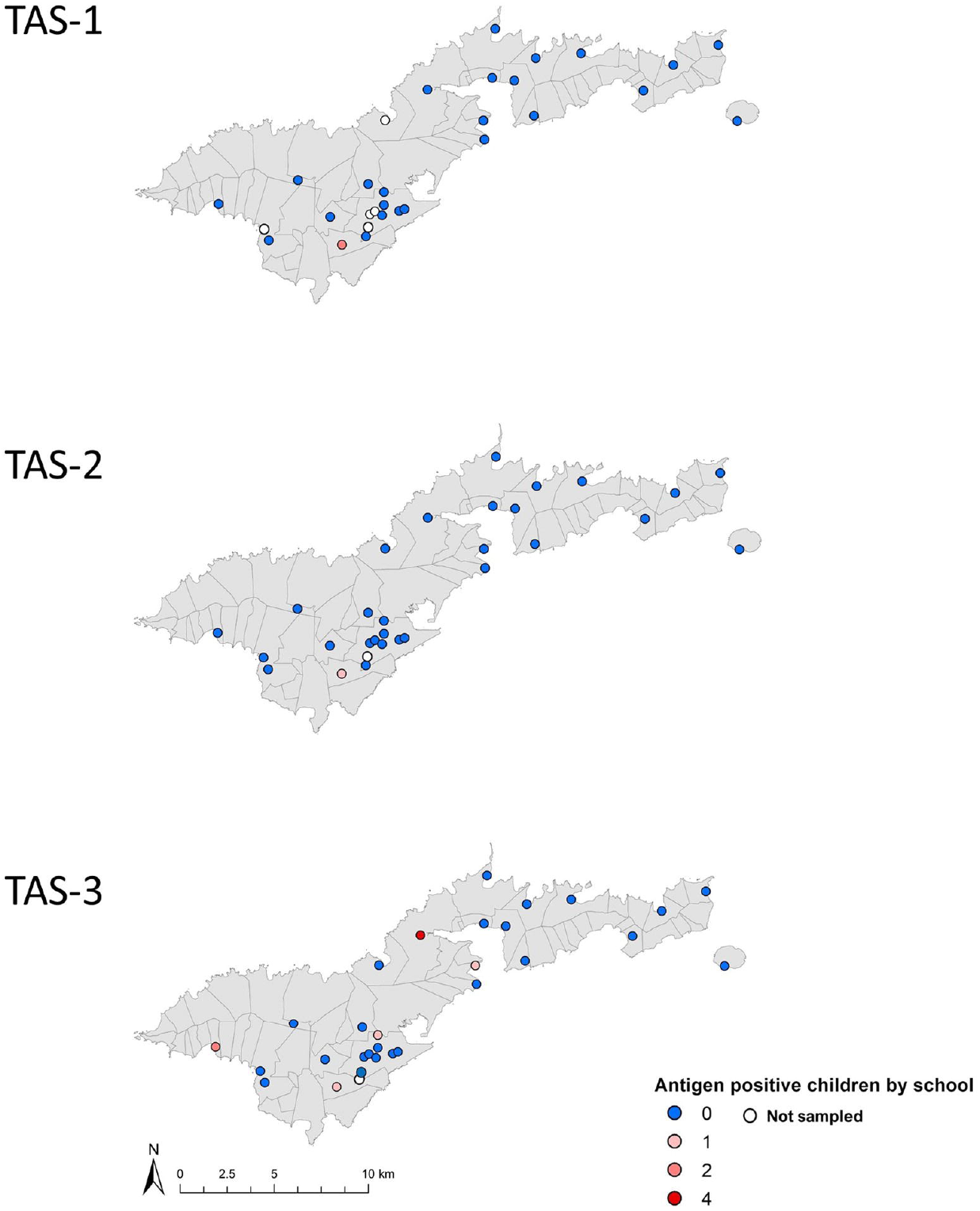
Locations of the schools (N=30) included in the surveys and observed number of antigen-positive children in TAS-1 (2011), TAS-2 (2015) and TAS-3 (2016) in American Samoa.

Supplementary table 2 shows the crude prevalence of Ab-positive children for each school in TAS-1, TAS-2 and TAS-3. In TAS-1, results of Ab responses were available for 1,112 schoolchildren; the highest overall Ab prevalence was observed for Bm33 (12.0%, 95% CI: 10.1%-14.0%), followed by Bm14 (6.8%, 95% CI: 5.4%-8.5%) and Wb123 (1.0%, 95% CI: 0.5%-17.6%). In TAS-2 and TAS-3, Ab results were available for 836 and 1,139 children, respectively; responses to Bm33 were also the highest in both surveys (7.8%, 95% CI: 6.1%-9.8% in TAS-2 and 20.8%, 95% CI:18.5%-23.3% in TAS-3). Figure 3 shows the school-level Ab prevalence for Wb123, Bm14 and Bm33 in each survey. In supplementary Table 3, the Ab status of all Ag-positive children is shown for each TAS. All Ag-positive children in TAS-1 and TAS-2 were seropositive for all three Abs. Six of the nine Ag-positive children in TAS-3 were seropositive for all Abs, and the remainder were seropositive for at least one Ab.

**Figure 3.**
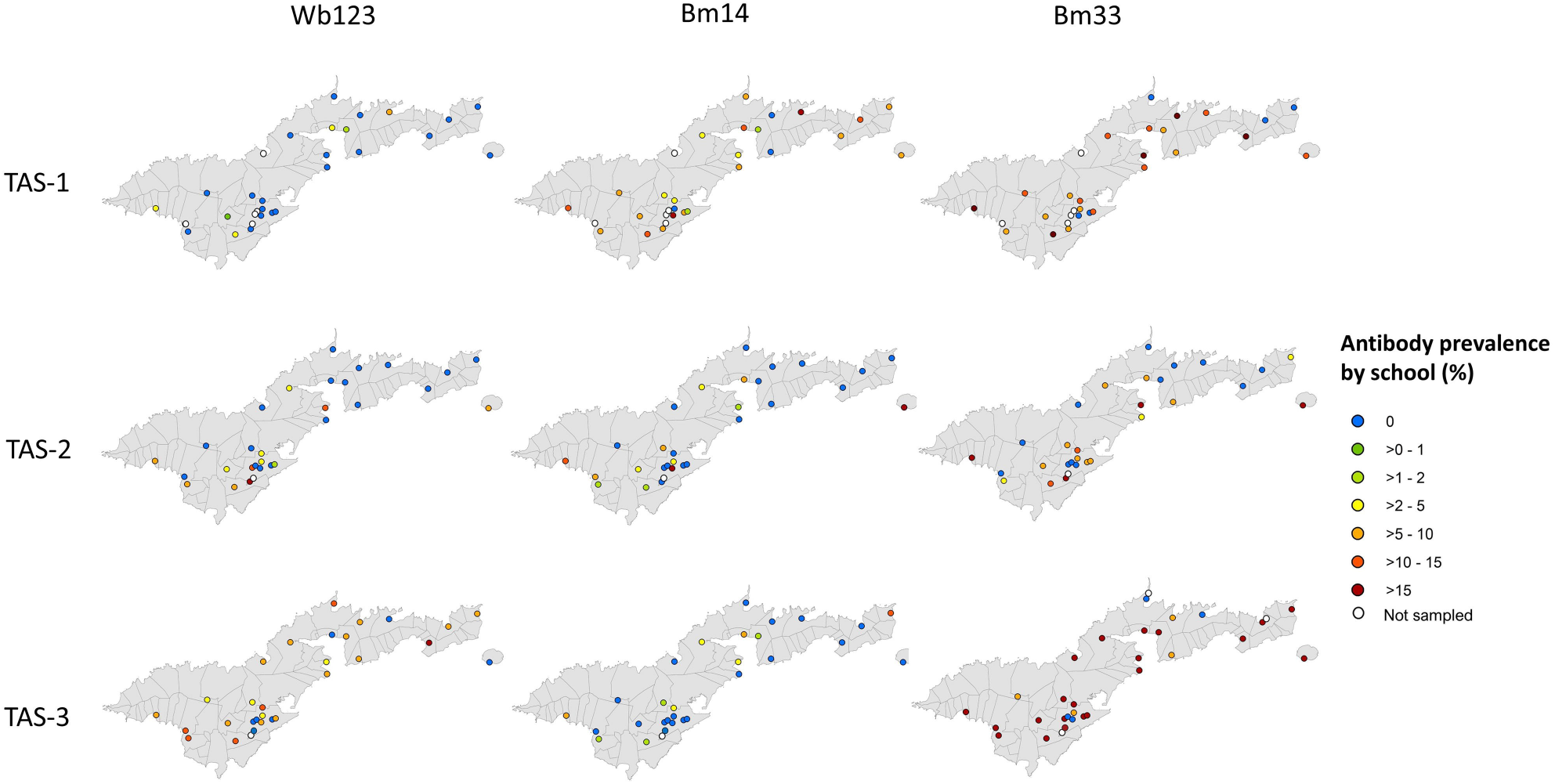
School locations (N=30) included in surveys and prevalence of antibody responses to Wb123, Bm14 and Bm33 in TAS-1 (2011), TAS-2 (2015) and TAS-3 (2016) in American Samoa.

Considering Ag and Ab status at the school level (presence or absence of Ag- and Ab-positive children), Figure 4 shows the percentage of schools that were Ag- and Ab-positive in TAS-1, TAS-2 and TAS-3 among all schools that participated in at least one TAS, and among the 25 schools that participated in all three TAS. The percentage of Bm14 Ab-positive schools decreased, while the percentage of Wb123 Ab-positive schools increased over time. Changes in percentage of schools that were positive for different combinations of Abs are also shown in Figure 3.

**Figure 4.**
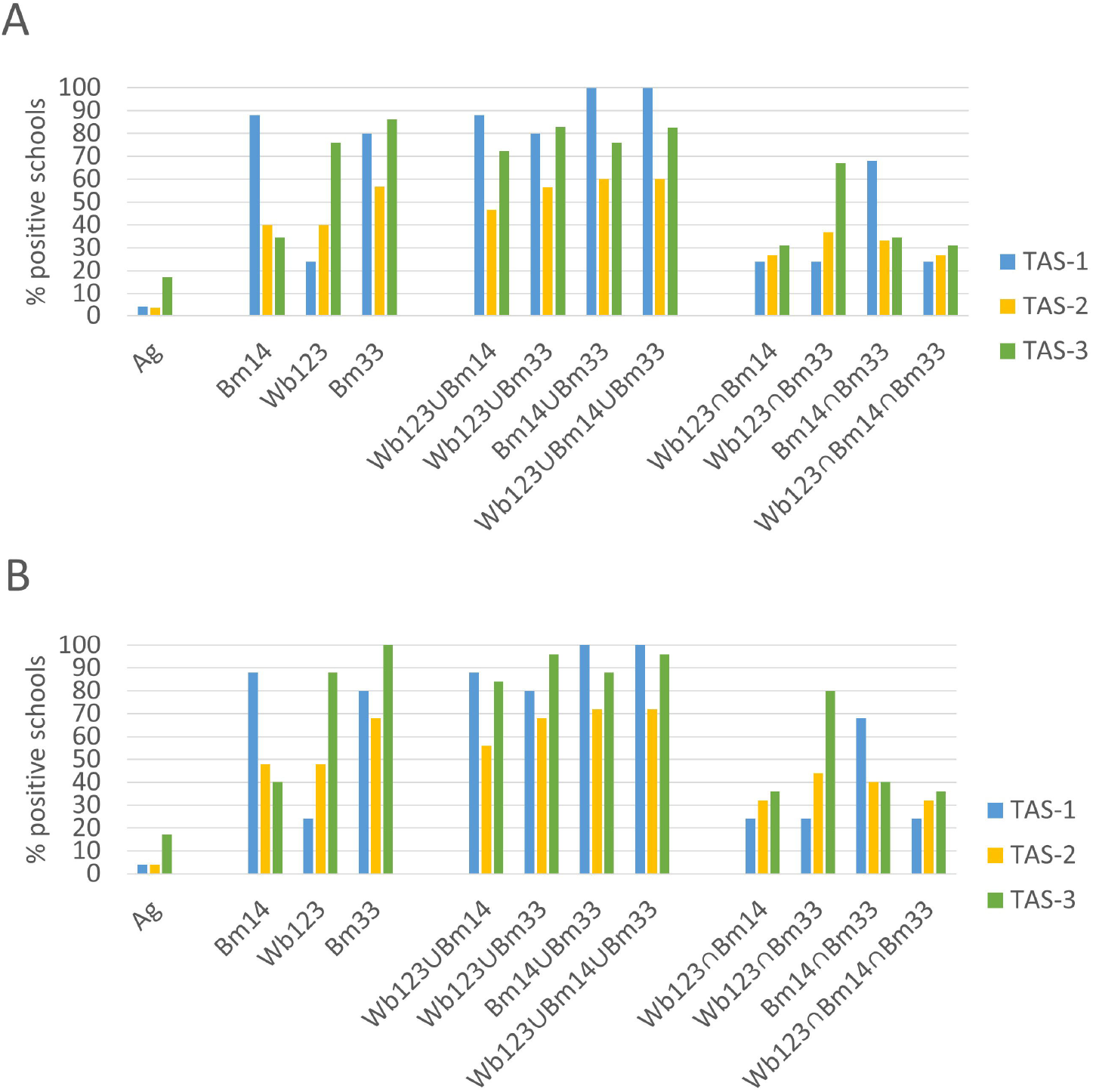
Percentage of Ag- and Ab-positive schools (including all combinations of Abs) among (A) all school locations (n=30) that participated in TAS-1, TAS-2 and TAS-3, and (B) the 25 schools that participated in all three TAS.

The percentage of Ab-positive children stratified by the school Ag status at TAS-3 was also examined. Figure 5 shows that in each TAS, Ab prevalence was higher in children who attended schools that were Ag-positive in TAS-3 compared to those who attended Ag-negative schools in TAS-3. In general, single Ab results in TAS-1 and Ag-negative schools in TAS-2 follow a similar trend with the highest prevalence for Bm33 Ab, followed by Bm14 and Wb123. A shift was observed in TAS-2 for schools that were Ag-positive in TAS-3, and all schools in TAS-3, towards higher prevalence of Wb123 than Bm14 Ab.

**Figure 5.**
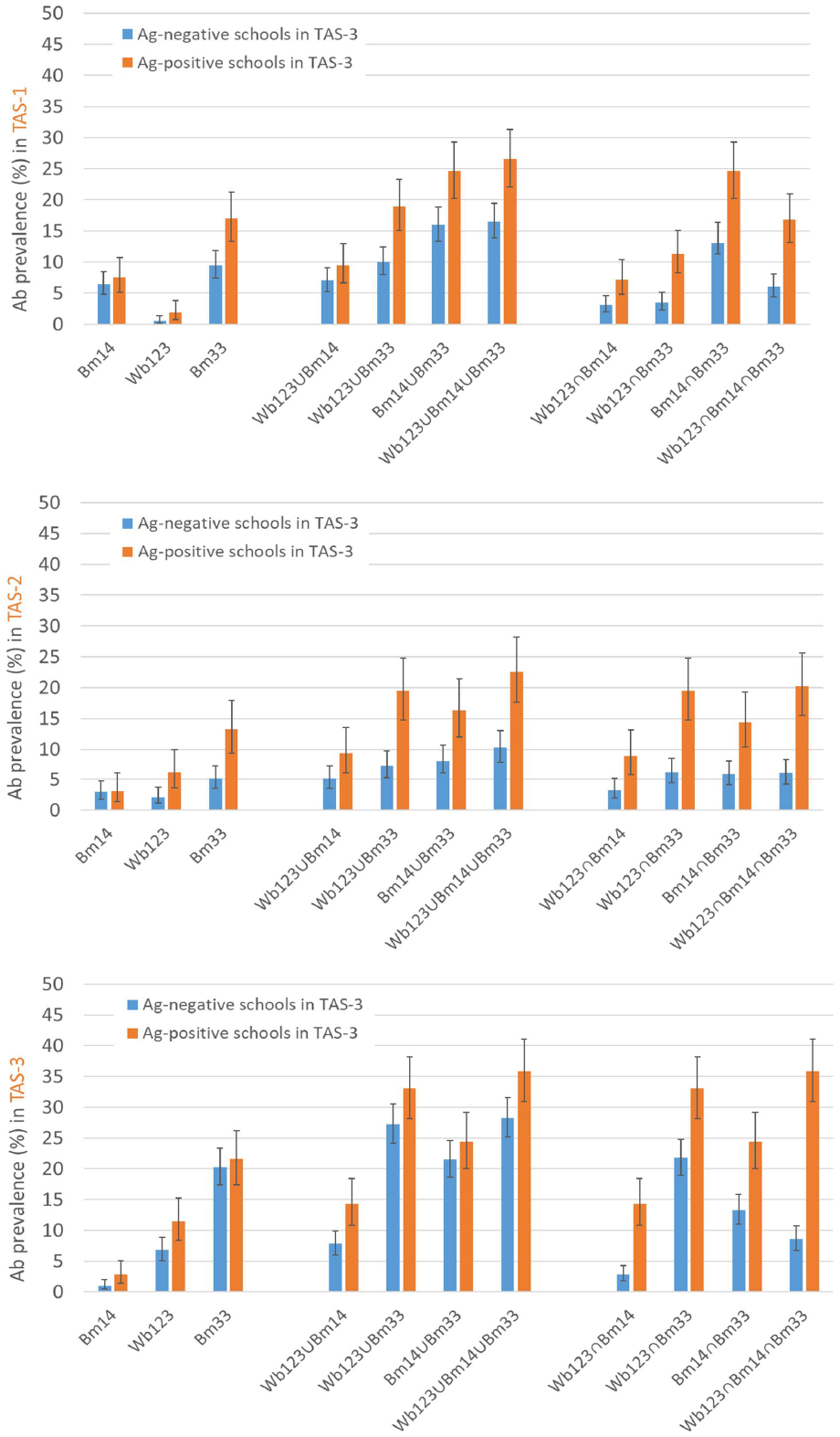
Crude prevalence of Ab-positive responses to single (Bm14, Bm33, Wb123) and combinations of Ab responses (Wb123∪Bm14, Wb123∪Bm33, Bm14∪Bm33, Wb123∪Bm14∪Bm33, Wb123∩Bm14, Wb123∩Bm33, Bm14∩Bm33, Wb123∩Bm14∩Bm33) in TAS-1, TAS-2 and TAS-3 among children from the 25 schools that participated in all three TAS, stratified by school Ag status (presence or absence of Ag-positive children) in TAS-3.

### Associations between school Ab status in TAS-1 and TAS-2 and school Ag status in TAS-3

To assess whether the school’s Ab-positive status at one survey time-point was associated with Ag-positive status in a later survey, we compared these seromarkers in TAS-3 and prior surveys. At the school level, significant statistical association was found between Wb123 Ab-positive status in TAS-2 and Ag-positive status in TAS-3 (Pearson’s Chi-Square value (X^2^: 5.36, df: 1, p-value: 0.02). Pearson’s chi-squared tests also show that some combinations of positive Abs (Bm14∪Bm33 and Wb123∪Bm14∪Bm33) in TAS-1 and TAS-2 were also statistically significantly associated with Ag-positive school status in TAS-3 (both with the same X^2^ value: 9.00, df: 1, p-value: 0.002). Table 2 shows the sensitivity, specificity, positive predictive value (PPV) and negative predictive value (NPV) for using school Ab status in TAS-1 and TAS-2 as indicators of school Ag status in TAS-3. Positive responses to Bm33, Bm14, Wb123∪Bm14, Wb123∪Bm33, Bm14∪Bm33, Wb123∪Bm14∪Bm33 and Bm14∩Bm33 in TAS-1, and all single and combinations of Ab responses in TAS-2 would have predicted Ag-positive schools in TAS-3 with high sensitivity (>80%) but low to moderate specificity (25% to 80%). From the positive Ab responses in TAS-2, Wb123∩Bm14 and Wb123∩Bm14∩Bm33 had the highest sensitivity (80%, 95% CI: 29%-99%) and specificity (80%, CI:56%-94%) results. The findings also revealed that Wb123 alone, Wb123∩Bm14, Wb123∩Bm33 and Wb123∩Bm14∩Bm33 in TAS-1 were less sensitive (40%, 95% CI:5%-85%) indicators of Ag-positive children in TAS-3, but more specific (80%, 95% CI: 56%-94%).

**Table 2.** Sensitivity, specificity, positive predictive value (PPV) and negative predictive value (NPV) for using school Ab status in TAS-1 and TAS-2 as indicators of school Ag.

| Indicator | School Ab status | Number of total Ab + schools in TAS-1 or TAS-2 | Number of schools Ab+ in TAS-1 or TAS-2 and Ag-positive in TAS-3 | Sensitivity % (95% CI) | Specificity % (95% CI) | PPV % (95% CI) | NPV % (95% CI) |
| --- | --- | --- | --- | --- | --- | --- | --- |
| Ab responses in TAS-1 as indicator of Ag-positive schools in TAS-3 | **Positive for single Ab** |  |  |  |  |  |  |
|  | Wb123 | 6 | 2 | 40 (5-85) | 80 (56-94) | 33 (4-78) | 84 (60-97) |
|  | Bm14 | 22 | 5 | 100 (48-100) | 15 (5-38) | 23 (8-45) | 100 (29-100) |
|  | Bm33 | 20 | 5 | 100 (48-100) | 25 (9-49) | 25 (9-49) | 100 (48-100) |
|  | **Positive for at least one Ab in the combination** |  |  |  |  |  |  |
|  | Wb123∪Bm14 | 22 | 5 | 100 (48-100) | 15 (3-38) | 23 (8-45) | 100 (29-100) |
|  | Wb123∪Bm33 | 20 | 5 | 100 (48-100) | 25 (9-49) | 25 (9-49) | 100 (48-100) |
|  | Bm14∪Bm33 | 25 | 5 | 100 | 0 | 20 | 0 |
|  | Wb123∪Bm14∪Bm33 | 25 | 5 | 100 | 0 | 20 | 0 |
|  | **Positive for all Abs in the combination** |  |  |  |  |  |  |
|  | Wb123∩Bm14 | 6 | 2 | 40 (5-85) | 80 (56-94) | 33 (4-78) | 84 (60-97) |
|  | Wb123∩Bm33 | 6 | 2 | 40 (5-85) | 80 (56-94) | 33 (4-78) | 84 (60-97) |
|  | Bm14∩Bm33 | 17 | 5 | 100 (48-100) | 40 (19-64) | 29 (10-56) | 100 (63-100) |
|  | Wb123∩Bm14∩Bm33 | 6 | 2 | 40 (5-85) | 80 (56-94) | 33 (4-78) | 84 (60-97) |
| Ab responses in TAS-2 as indicators of Ag-positive schools in TAS-3 | **Positive for single Ab** |  |  |  |  |  |  |
|  | Wb123 | 11 | 5 | 100 (48-100) | 70 (46-88) | 45 (17-77) | 100 (77-100) |
|  | Bm14 | 11 | 4 | 80 (28-99) | 65 (41-85) | 36 (11-69) | 93 (66-100) |
|  | Bm33 | 17 | 5 | 100 (48-100) | 40 (19-64) | 29 (10-56) | 100 (63-100) |
|  | **Positive for at least one Ab in the combination** |  |  |  |  |  |  |
|  | Wb123∪Bm14 | 14 | 5 | 100 (48-100) | 55 (32-77) | 36 (13-65) | 100 (72-100) |
|  | Wb123∪Bm33 | 17 | 5 | 100 (48-100) | 40 (19-64) | 29 (10-56) | 100 (63-100) |
|  | Bm14∪Bm33 | 18 | 5 | 100 (48-100) | 35 (15-59) | 28 (10-53) | 100 (59-100) |
|  | Wb123∪Bm14∪Bm33 | 18 | 5 | 100 (48-100) | 35 (15-59) | 28 (10-53) | 100 (59-100) |
|  | **Positive for all Abs in the combination** |  |  |  |  |  |  |
|  | Wb123∩Bm14 | 8 | 4 | 80 (28-99) | 80 (56-94) | 50 (16-84) | 94 (71-100) |
|  | Wb123∩Bm33 | 11 | 5 | 100 (48-100) | 70 (46-88) | 45 (17-77) | 100 (77-100) |
|  | Bm14∩Bm33 | 10 | 4 | 80 (28-99) | 70 (46-88) | 40 (12-74) | 93 (68-100) |
|  | Wb123∩Bm14∩Bm33 | 8 | 4 | 80 (28-99) | 80 (56-94) | 50 (16-84) | 94 (71-100) |

### Prediction of school Ag status in TAS-3 based on school Ab status in TAS-1 and TAS-2

The results of the regression analyses (Table 3) indicate that Wb123 Ab-positive schools in TAS-2 were significantly associated with Ag-positive status in TAS-3 (odds ratio (OR): 24.5, 95% CI:1.17-512.6). Similarly, schools that were positive for Wb123∩Bm14, Wb123∩Bm33 and Wb123∩Bm14∩Bm33 in TAS-2 also had higher odds of being Ag-positive in TAS-3 (OR ranging from 16.0 to 24.5). Schools that were Ab-positive for Bm14∪Bm33 and Wb123∪Bm14∪Bm33 in TAS-1 also had significantly higher odds of being Ag-positive in TAS-3.

**Table 3.** Antibody positivity (at school level) in TAS-1 and TAS-2 as predictors of school Ag status in TAS-3.

|  | Predictors | Number of Ag-positive schools in TAS-3 | OR | 95% CI |
| --- | --- | --- | --- | --- |
| TAS-1 | **Positive for individual Ab** |  |  |  |
|  | Wb123 | 2 | 2.6 | 0.3-21.7 |
|  | Bm14 | 5 | 2.2 * | 0.1-49.5 |
|  | Bm33 | 5 | 3.9 * | 0.2-82.8 |
|  | **Positive for at least one Ab in the combination** |  |  |  |
|  | Wb123∪Bm14 | 5 | 2.2 * | 0.1-49.5 |
|  | Wb123∪Bm33 | 5 | 3.9 * | 0.2-82.8 |
|  | Bm14∪Bm33 | 5 | 451 * | 0.8-25,409 |
|  | Wb123∪Bm14∪Bm33 | 5 | 451 * | 0.8-25,409 |
|  | **Positive for all Abs in the combination** |  |  |  |
|  | Wb123∩Bm14 | 5 | 2.6 | 0.3-21.7 |
|  | Wb123∩Bm33 | 5 | 2.6 | 0.3-21.7 |
|  | Bm14∩Bm33 | 5 | 7.4 * | 0.4-153.8 |
|  | Wb123∩Bm14∩Bm33 | 5 | 2.6 | 0.3-21.7 |
| TAS-2 | **Positive for individual Ab** |  |  |  |
|  | Wb123 Ab | 5 | **24.5 \*** | **1.2-512.7** |
|  | Bm14 Ab | 4 | 7.4 | 0.7-80.0 |
|  | Bm33 Ab | 5 | 7.5 * | 0.4-143.8 |
|  | **Positive for at least one Ab in the combination** |  |  |  |
|  | Wb123∪Bm14 | 5 | 13.3 * | 0.7-272.8 |
|  | Wb123∪Bm33 | 5 | 7.5 * | 0.4-153.8 |
|  | Bm14∪Bm33 | 5 | 2.6 * | 0.3-21.7 |
|  | Wb123∪Bm14∪Bm33 | 5 | 2.6 * | 0.3-21.7 |
|  | **Positive for all Abs in the combination** |  |  |  |
|  | Wb123∩Bm14 | 5 | **16.0** | **1.4-185.4** |
|  | Wb123∩Bm33 | 5 | **24.5 \*** | **1.2-512.6** |
|  | Bm14∩Bm33 | 5 | 2.7 | 0.3-21.7 |
|  | Wb123∩Bm14∩Bm33 | 5 | **16.0** | **1.4-185.4** |

\*Note that accurate ORs could not be calculated because all Ab-positive schools in these categories were Ag-positive in TAS-3. Reported ORs were calculated after applying Haldane's correction.

## Discussion

Our study compared Ag and Ab results obtained in the three TAS conducted in American Samoa in 2011, 2015 and 2016. We found that the school Ab status in TAS-1 and TAS-2 was significantly associated with the presence of Ag-positive children in TAS-3 and could have provided an earlier indication of resurgence than the use of Ag alone. The results suggest that anti-filarial Ab responses among young children may be used as early signals of ongoing transmission or resurgence in a post-MDA setting. The findings also showed that the statistically significant associations between responses to Wb123, Wb123∩Bm14 and Wb123∩Bm14∩Bm33, at the school level in TAS-2 and Ag-positive schools in TAS-3 provided the best balance of sensitivity (80%) and specificity (80%) test results.

The serological patterns of the Ab responses to Wb123, Bm14 and Bm33 Ags varied across the three TAS. The overall and school-level prevalence of Bm33 Ab was the highest in all TAS. Also, Bm33 Ab and all sets of combinations that included this Ab were highly prevalent in both Ag-positive and negative schools in TAS-3. These findings concur with previous studies that found Bm33 Ab as the first detectable seromarker that induces an Ab response even without high levels of antigenemia (Hamlin et al., 2012). In contrast, the prevalence of Wb123 Ab increased from TAS-1 to TAS-3. The kinetics of Wb123Ab are not currently well understood. It has been proposed that the response to Wb123 is developed after repeated larval stimulation which is then required to sustain an Ab response to Wb123 (Kubofcik et al., 2012). Therefore, it is expected that as infection rates increase, the prevalence of Wb123 responses will follow the same trend. The decrease in prevalence of Bm14 Ab over the three TAS was an unexpected serological pattern that differed from previous reports (Hamlin et al., 2012). The discordant results between Ag and the three Abs suggests that serological responses in recently acquired infections is complex and that further studies are required to fully understand and interpret Ag and Ab profiles (Moss et al., 2011). In pre-MDA settings, it is expected that Ab responses are concordant as there is no variability in the intensity of Ag exposure (Hamlin et al., 2012). After MDA interventions, studies conducted in different settings have shown that Ab responses tend to decline (Ramzy et al., 2006, Tisch et al., 2008). Ab against Bm14 and Bm33 decreased significantly in Ag-negative children treated with diethylcarbamazine in Haiti (Moss et al., 2011). Also, in Egypt and Papua New Guinea, observations showed that positive responses to Bm14 converted to negative in individuals treated with anti-filarial drugs. Currently, there is little evidence on the timing and magnitude of Ag and Ab responses after infection (Moss et al., 2011).

Increasing evidence indicates that the use of Ag alone in TAS may not be sufficiently sensitive for making decisions to stop MDA or for post-MDA surveillance (Gass et al., 2012, Hamlin et al., 2012, Won Kimberly Y et al., 2018). Although Sri Lanka was recognised as having eliminated LF as a public health problem in 2016, more recent studies found low-level persistence of infection in some regions (Rao et al., 2018). Similarly, other LF-endemic areas including Tonga, American Samoa and India, which had utilised school-based or community TAS for post-MDA surveillance also found that TAS alone was not sufficiently sensitive for programmatic decision-making (Joseph Hayley et al., 2011, Lau et al., 2020, Subramanian et al., 2020).

In the context of American Samoa, while the territory passed TAS-1 and TAS-2, the surveys failed to detect hotspots and residual ongoing transmission, resulting in resurgence of LF and ultimately failing TAS-3 (Sheel et al., 2018). Therefore, alternative surveillance methods are required to improve the prompt identification of ongoing transmission or resurgence in low prevalence settings. This is particularly important in the post-MDA setting when residual infections can be highly spatially heterogenous (Lau et al., 2014).

As initiatives to incorporate Ab testing as a tool to strengthen post-MDA surveillance have increasingly been proposed, additional work is needed to assess the performance of Ab assays and potential limitations of using Ab testing to complement TAS. This is particularly important in areas of co-endemicity with other filarial infections, where cross-reactivity with Ags from other filarial parasites such as *Onchocerca volvulus* and *Loa loa* have been documented (Dolo et al., 2019, Hertz et al., 2018).

The limitations of this study include the use of different Ag tests and Ab assays over the three TAS. The use of different cut-off values to define Ag- or Ab-positivity also pose challenges for the interpretation of results. Therefore, further work is needed to standardise tests, and revise and propose consistent cut-off thresholds for multibead assays. Longitudinal data on individuals that allow us to examine the timing and magnitude of Ag and Ab responses and document immunological profile to LF infection is currently unavailable but would be important for assessing Ag and Ab trends over time. At the time of TAS-1, LIPS was the only option available for including Wb123 and samples from TAS-1 and TAS-2 were not repeated when TAS-3 was completed. Unfortunately, it is not always feasible to repeat testing of samples from large studies or to perform concurrent testing from multiple surveys conducted at different times. Therefore, the inability to retest samples and to use a consistent platform may be a limitation for the analysis of the Wb123 responses.

This study provides important evidence that helps better understand anti-filarial antibodies in children in the context of LF resurgence. The associations found between the school Ab status in TAS-1 and TAS-2 and the school Ag status in TAS-3 suggests that Abs could have provided an earlier indication of LF resurgence in American Samoa. Although our study was conducted on data from American Samoa, the concepts are widely applicable to other settings globally. Our findings contribute new evidence for the potential role of Ab testing, as an additional monitoring tool, in guiding programmatic decision making, strengthening post-MDA surveillance, and optimising successful elimination of LF. Further studies are required to better understand specific Ab responses after MDA.

## Data Availability

The data used in the present study are available from the corresponding author on reasonable request.

## Contributors

AMC, CLL and PMG developed the study conception and design. Analyses were performed by AMCR and CLL. AMCR and CLL drafted the manuscript. All authors helped in the interpretation of results and critically reviewed the manuscript.

## Declaration of interests

We declare no competing interests.

## Patient consent for publication

Not required

## Acknowledgements

This study was supported by the Coalition for Operational Research on Neglected Tropical Diseases (COR-NTD), which is funded at The Task Force for Global Health primarily by the Bill & Melinda Gates Foundation [OPP1053230], the United Kingdom Department for International Development, and by the United States Agency for International Development through its Neglected Tropical Diseases Program. CLL was supported by Australian National Health and Medical Research Council Fellowships (APP1193826). MS is supported by a fellowship from the Westpac Scholars Trust.

## Notes

### Competing Interest Statement

The authors have declared no competing interest.

### Author Declarations

Ethics approvals for TAS-1 and TAS-2 were granted by the DOH Institutional Review Board (IRB) and the U.S. Centers for Disease Control and Prevention (CDC) as program evaluation, non-research. (Won Kimberly Y. et al., 2018). TAS-3 was approved by the American Samoa Institutional Review Board and the Human Research Ethics Committee at the Australian National University (protocol number 2016/482).

