## Supplementary tables for "Potential use of antibodies to provide an earlier indication of lymphatic filariasis resurgence in post-mass drug administration surveillance, American Samoa"

**Supplementary table 1 Crude Ag prevalence and 95% confidence intervals for each school in TAS-1 (2011), TAS-2 (2015) and TAS-3 (2016) in American Samoa.**

|  | **TAS 1** | | | **TAS 2** | | | **TAS 3** | | |
| --- | --- | --- | --- | --- | --- | --- | --- | --- | --- |
| **School name** | **Ag positive/ total tested** | **Crude Prevalence**  **(%)** | **95% CI**  **(%)** | **Ag positive/ total tested** | **Crude prevalence**  **(%)** | **95% CI**  **(%)** | **Ag positive/ total tested** | **Crude Prevalence**  **(%)** | **95% CI**  **(%)** |
| **Afonotele** | 0/12 | 0.0 | 0.0-26.5 | 0/6 | 0.0 | 0.0-45.9 | 0/11 | 0.0 | 0.0-28.5 |
| **Alataua-Lua** | 0/38 | 0.0 | 0.0-9.3 | 0/29 | 0.0 | 0.0-11.9 | 2/44 | 4.5 | 0.5-15.5 |
| **Alofau** | 0/18 | 0.0 | 0.0-18.5 | 0/13 | 0.0 | 0.0-24.7 | 0/12 | 0.0 | 0.0-26.5 |
| **Aua** | 0/47 | 0.0 | 0.0-7.6 | 0/14 | 0.0 | 0.0-23.2 | 0/65 | 0.0 | 0.0-5.5 |
| **A. P. Lutali (Aunuu)** | 0/16 | 0.0 | 0.0-20.6 | 0/12 | 0.0 | 0.0-26.5 | 0/9 | 0.0 | 0.0-33.6 |
| **Coleman (Pago)** | 0/63 | 0.0 | 0.0-5.7 | 0/37 | 0.0 | 0.0-9.5 | 4/82 | 4.9 | 1.3-12.0 |
| **Kanana Fou** | 0/10 | 0.0 | 0.0-30.8 | 0/40 | 0.0 | 0.0-8.8 | 0/29 | 0.0 | 0.0-11.9 |
| **Laulii** | 0/25 | 0.0 | 0.0-13.7 | 0/18 | 0.0 | 0.0-18.5 | 0/31 | 0.0 | 0.0-11.2 |
| **Le’atele (Fagasa)** | - | - | - | 0/4 | 0.0 | 0.0-60.2 | 0/13 | 0.0 | 0.0-24.7 |
| **Leone Midkiff** | 0/113 | 0.0 | 0.0-3.2 | 0/76 | 0 | 0.0-4.7 | 0/59 | 0.0 | 0.0-6.1 |
| **Lupelele** | 2/92 | 2.2 | 0.3-7.6 | 1/85 | 1.2 | 0.0-6.4 | 1/94 | 1.1 | 0.0-5.8 |
| **Manulele** | 0/90 | 0.0 | 0.0-4.0 | 0/44 | 0.0 | 0.0-8.0 | 1/93 | 1.1 | 0.0-5.8 |
| **Manumalo Baptist School** | 0/48 | 0.0 | 0.0-7.4 | 0/44 | 0.0 | 0.0-8.0 | 0/72 | 0.0 | 0.0-5.0 |
| **Masefau** | 0/13 | 0.0 | 0.0-24.7 | 0/9 | 0.0 | 0.0-33.6 | 0/14 | 0.0 | 0.0-23.2 |
| **Matafao** | 0/34 | 0.0 | 0.0-10.3 | 0/35 | 0.0 | 0.0-10.0 | 1/44 | 2.3 | 0.1-12.0 |
| **Matatula** | 0/14 | 0.0 | 0.0-23.2 | 0/26 | 0.0 | 0.0-13.2 | 0/27 | 0.0 | 0.0-12.7 |
| **Mt. Alava (Vatia)** | 0/11 | 0.0 | 0.0-28.5 | 0/10 | 0.0 | 0.0-30.8 | 0/9 | 0.0 | 0.0-33.6 |
| **Olomoana (Aoa)** | 0/26 | 0.0 | 0.0-13.2 | 0/6 | 0.0 | 0.0-45.9 | 0/22 | 0.0 | 0.0-15.4 |
| **Pacific Horizon** | - | - | - | 0/1 | 0.0 | 0.0-97.5 | 0/4 | 0.0 | 0.0-60.2 |
| **Pavaiai** | 0/107 | 0.0 | 0.0-3.4 | 0/89 | 0.0 | 0.0-4.1 | 0/162 | 0.0 | 0.0-2.2 |
| **Peteli Academy** | - | - | - | 0/8 | 0.0 | 0.0-36.9 | 0/15 | 0.0 | 0.0-21.8 |
| **Samoa Baptist Academy** | 0/3 | 0.0 | 0.0-70.8 | 0/12 | 0.0 | 0.0-26.5 | 0/17 | 0.0 | 0.0-19.5 |
| **(Iakina Adventist Academy) SDA** | 0/11 | 0.0 | 0.0-28.5 | 0/10 | 0.0 | 0.0-30.8 | 0/9 | 0.0 | 0.0-33.6 |
| **Siliaga** | 0/28 | 0.0 | 0.0-12.3 | 0/14 | 0.0 | 0.0-23.2 | 0/34 | 0.0 | 0.0-10.3 |
| **South Pacific Academy** | 0/7 | 0.0 | 0.0-40.9 | 0/9 | 0.0 | 0.0-33.6 | 0/12 | 0.0 | 0.0-26.5 |
| **South Pacific International Christian Center** | 0/25 | 0.0 | 0.0-13.7 | 0/27 | 0.0 | 0.0-12.8 | 0/30 | 0.0 | 0.0-11.6 |
| **St. Francis** | 0/28 | 0.0 | 0.0-12.3 | 0/10 | 0.0 | 0.0-30.8 | 0/13 | 0.0 | 0.0-24.7 |
| **St. Theresa** | 0/0 | 0.0 | 0.0-100.0 | 0/18 | 0.0 | 0.0-18.5 | 0/20 | 0.0 | 0.0-16.8 |
| **Ta’iala Academy** | - | - | - | 0/2 | 0.0 | 0.0-84.2 | - | - | - |
| **Tafuna** | 0/57 | 0.0 | 0.0-6.3 | 0/60 | 0.0 | 0.0-6.0 | 0/97 | 0.0 | 0.0-3.7 |
| **Total** | **2/937** | **0.2** | **0.0-0.8** | **1/768** | **0.1** | **0.0-0.7** | **9/1,143** | **0.8** | **0.4-1.5** |

*The colour blue highlights schools with Ag-positive children

**Supplementary table 2 Crude prevalence and 95% confidence intervals of Ab responses to Wb123, Bm14 and Bm33 for each school in TAS-1 (2011), TAS-2 (2015) and TAS-3 (2016) in American Samoa. The colour blue highlights schools with Ab-positive children**

|  | **Wb123** | | | **Bm14** | | | **Bm33** | | |
| --- | --- | --- | --- | --- | --- | --- | --- | --- | --- |
| **School name** | **TAS-1**  **%**  **(95%CI)** | **TAS-2**  **%**  **(95%CI)** | **TAS-3**  **%**  **(95%CI)** | **TAS-1**  **%**  **(95%CI)** | **TAS-2**  **%**  **(95%CI)** | **TAS-3**  **%**  **(95%CI)** | **TAS-1**  **%**  **(95%CI)** | **TAS-2**  **%**  **(95%CI)** | **TAS-3**  **%**  **(95%CI)** |
| **Afonotele** | 0.0  (0-21.8) | 0.0  (0.0-52.2) | 9.1  (0.2-41.3) | 0.0  (0-21.8) | 0.0  (0.0-52.2) | 0.0  (0.0-28.5) | 26.7  (7.8-55.1) | 0.0  (0.0-52.2) | 9.1  (0.2-41.3) |
| **Alataua-Lua** | 5.0  (0.6-16.9) | 9.7  (2.0-25.7) | 22.7  (11.5-37.8) | 12.5  (4.2-26.8) | 12.9  (3.6-29.8) | 6.8  (1.4-18.7) | 20.0  (9.1-35.6) | 19.4  (7.5-37.5) | 27.3  (15.0-42.8) |
| **Alofau** | 0.0  (0-16.1) | 0.0  (0.0-20.6) | 16.7  (2.1-48.4) | 8.3  (1.1-27.0) | 0.0  (0.0-20.6) | 0.0  (0.0-26.5) | 20.8  (7.1-42.1) | 0.0  (0.0-20.6) | 25.0  (5.5-57.2) |
| **Aua** | 1.8  (0-9.5) | 0.0  (0.0-13.7) | 6.2  (1.7-15.0) | 1.8  (0.1-9.5) | 0.0  (0.0-13.7) | 1.5  (0.0-8.3) | 5.3  (1.1-14.9) | 0.0  (0.0-13.7) | 16.9  (8.8-28.3) |
| **A. P. Lutali (Aunuu)** | 0.0  (0-18.5) | 9.1  (0.2-41.3) | 0.0  (0.0-33.6) | 5.5  (0.1-27.3) | 18.2  (2.3-51.8) | 0.0  (0.0-33.6) | 11.1  (1.4-34.7) | 18.2  (2.3-51.8) | 22.2  (2.8-60.0) |
| **Coleman (Pago)** | 0.0  (0-5.1) | 2.5  (0.1-13.1) | 9.8  (4.3-18.3) | 2.8  (0.3-9.9) | 5.0  (0.6-16.9) | 2.4  (0.3-8.5) | 11.4  (5.1-21.3) | 7.5  (1.6-20.4) | 19.5  (11.6-29.7) |
| **Iakina Adventist Academy (SDA)** | 0.0  (0-21.8) | 16.7  (2.1-48.4) | 0.0  (0.0-33.6) | 6.7  (0.2-31.9) | 0.0  (0.0-26.5) | 0.0  (0.0-33.6) | 6.7  (0.2-31.9) | 16.7  (2.1-48.4) | 22.2  (2.8-60.0) |
| **Kanana Fou** | 0.0  (0-30.8) | 2.4  (0.1-12.8) | 3.4  (0.1-17.8) | 0.0  (0-30.8) | 4.9  (0.6-16.5) | 0.0  (0.0-11.9) | 10.0  (0.2-44.5) | 7.3  (1.5-19.9) | 6.9  (0.8-22.8) |
| **Lauli’i** | 0.0  (0-11.6) | 0.0  (0.0-18.5) | 10.0  (2.1-26.5) | 0.0  (0-11.6) | 0.0  (0.0-18.5) | 0.0  (0.0-11.6) | 10.0  (2.1-26.5) | 5.6  (0.1-27.3) | 10.0  (2.1-26.5) |
| **Le’atele (Fagasa)** | - | 0.0  (0.0-30.8) | 7.7  (0.2-36.0) | - | 0.0  (0.0-30.8) | 0.0  (0.0-24.7) | - | 0.0  (0.0-30.8) | 30.8  (9.1-61.4) |
| **Leone Midkiff** | 0.0  (0-2.8) | 5.3  (1.5-13.1) | 10.5  (4.0-21.5) | 7.7  (3.8-13.8) | 1.3  (0.0-7.2) | 1.7  (0.0-9.4) | 9.3  (4.9-15.7) | 2.7  (0.3-9.3) | 17.5  (8.7-29.9) |
| **Lupelele** | 4.8  (1.6-10.8) | 5.9  (1.9-13.2) | 11.7  (6.0-20.0) | 14.3  (8.2-22.5) | 1.2  (0.0-6.4) | 1.1  (0.0-5.8) | 25.7  (17.7-35.2) | 12.9  (6.6-22.0) | 10.0  (9.2-25.0) |
| **Manulele** | 0.0  (0-3.1) | 2.1  (0.1-11.1) | 10.8  (5.3-18.9) | 4.2  (1.4-9.6) | 0.0  (0.0-7.4) | 3.2  (0.7-9.1) | 11.0  (6.0-18.1) | 10.4  (3.5-22.7) | 28.0  (19.1-38.2) |
| **Manumalo Baptist School** | 0.0  (0-5.8) | 0.0  (0.0-7.9) | 4.2  (0.9-11.7) | 3.2  (0.4-11.2) | 8.9  (2.5-21.2) | 1.4  (0.0-7.5) | 8.1  (2.7-17.8) | 6.7  (1.4-18.3) | 19.4  (11.1-30.5) |
| **Masefau** | 6.2  (0.2-30.2) | 0.0  (0.0-26.5) | 0.0  (0.0-23.2) | 18.7  (4.0-45.6) | 0.0  (0.0-26.5) | 0.0  (0.0-23.2) | 12.5  (1.5-38.3) | 0.0  (0.0-26.5) | 0.0  (0.0-23.2) |
| **Matafao** | 0.0  (0-9.5) | 11.3  (4.3-23.0) | 4.5  (0.6-15.5) | 2.7  (0.1-14.2) | 1.9  (0.0-10.1) | 2.3  (0.1-12.0) | 18.9  (8.0-35.1) | 17.0  (8.1-29.8) | 18.2  (8.2-32.7) |
| **Matatula** | 0.0  (0-23.2) | 0.0  (0.0-13.7) | 7.4  (0.9-24.3) | 7.1  (0.2-33.9) | 0.0  (0.0-13.7) | 14.8  (4.2-33.7) | 0.0  (0-23.2) | 4.0  (0.1-20.4) | 37.0  (19.4-57.6) |
| **Mt. Alava (Vatia)** | 0.0  (0-24.7) | 0.0  (0.0-26.5) | 11.1  (0.3-48.2) | 7.7  (0.2-36.0) | 0.0  (0.0-26.5) | 0.0  (0.0-33.6) | 0.0  (0-24.7) | 0.0  (0.0-26.5) | 0.0  (0.0-33.6) |
| **Olomoana (Aoa)** | 0.0  (0-14.2) | 0.0  (0.0-40.9) | 9.1  (1.1-29.2) | 14.8  (4.2-33.7) | 0.0  (0.0-40.9) | 0.0  (0.0-15.4) | 0  (0-12.8) | 0.0  (0.0-40.9) | 31.8  (13.9-54.9) |
| **Pacific Horizon** | - | 0.0  (0.0-97.5) | 0.0  (0.0-60.2) | - | 0.0  (0.0-97.5) | 0.0  (0.0-60.2) | - | 0.0  (0.0-97.5) | 0.0  (0.0-60.2) |
| **Pavaiai** | 0.6  (0-3.5) | 4.2  (1.2-10.5) | 8.1  (4.4-13.4) | 7.1  (3.6-12.3) | 4.3  (1.2-10.5) | 0.0  (0.0-2.3) | 8.4  (4.5-13.9) | 9.6  (4.5-17.4) | 30.4  (23.4-38.2) |
| **Peteli Academy** | - | 14.3  (0.4-57.9) | 0.0  (0.0-21.8) | - | 0.0  (0.0-40.9) | 0.0  (0.0-21.8) | - | 0.0  (0.0-40.9) | 26.7  (7.8-55.1) |
| **Samoa Baptist Academy** | 0.0  (0-60.2) | 0.0  (0.0-26.5) | 5.9  (0.1-28.7) | 25.0  (0.6-80.6) | 16.7  (2.1-48.4) | 0.0  (0.0-19.5) | 0.0  (0-60.2) | 0.0  (0.0-26.5) | 0.0  (0.0-19.5) |
| **Siliaga** | 0.0  (0-11.9) | 0.0  (0.0-20.6) | 2.9  (0.1-15.3) | 6.9  (0.8-22.8) | 0.0  (0.0-20.6) | 0.0  (0.0-10.3) | 10.3  (2.2-27.3) | 0.0  (0.0-20.6) | 8.8  (1.9-23.7) |
| **South Pacific Academy** | 0.0  (0-30.8) | 0.0  (0.0-24.7) | 0.0  (0.0-26.5) | 10.0  (0.2-44.5) | 0.0  (0.0-24.7) | 0.0  (0.0-26.5) | 0.0  (0-30.8) | 7.7  (0.2-36.0) | 16.7  (0.2-38.5) |
| **South Pacific International Christian Center** | 0.0  (0-11.9) | 0.0  (0.0-12.8) | 6.7  (0.8-22.1) | 6.9  (0.8-22.8) | 0  (0.0-12.7) | 0.0  (0.0-11.6) | 13.8  (3.9-31.7) | 3.7  (0.1-19.0) | 16.7  (5.6-34.7) |
| **St. Francis** | 3.6  (0.1-18.3) | 0.0  (0.0-30.8) | 0.0  (0.0-24.7) | 14.3  (4.0-32.7) | 10.0  (0.3-44.5) | 7.7  (0.2-36.0) | 14.3  (4.0-32.7) | 10.0  (0.3-44.5) | 30.8  (9.1-61.4) |
| **St. Theresa** | - | 0.0  (0.0-18.5) | 15.0  (3.2-37.9) | - | 5.6  (0.1-27.3) | 0.0.  (0.0-16.8) | - | 0.0  (0.0-18.5) | 20.0  (5.7-43.7) |
| **Ta’iala Academy** | - | 0.0  (0.0-84.2) | - | - | 0.0  (0.0-84.2) | - | - | 0.0  (0.0-84.2) | - |
| **Tafuna** | 0.0  (0-6.1) | 1.5  (0.0-8.3) | 7.2  (3.0-14.3) | 1.7  (0.1-9.2) | 0.0  (0.0-5.5) | 0.0  (0.0-3.7) | 13.8  (6.1-25.4) | 7.7  (2.5-17.0) | 20.6  (13.1-30.0) |
| **Overall Prevalence** | **1.0**  **(0.5-17.6)** | **3.6**  **(2.4-5.1)** | **8.3**  **(6.7-10.0)** | **6.8**  **(5.4-8.5)** | **3.0**  **(2.0-4.4)** | **1.6**  **(0.9-2.5)** | **12.0**  **(10.1-14.0)** | **7.8**  **(6.1-9.8)** | **20.8**  **(18.5-23.3)** |

**Supplementary table 3 Antibody status (Wb123, Bm14, Bm33) of antigen-positive children detected in TAS-1, TAS-2, and TAS-3 in American Samoa.**

| **TAS and school** | **Ag status** | **Wb123 Ab status** | **Bm14 Ab status** | **Bm33 Ab status** |
| --- | --- | --- | --- | --- |
| **TAS-1** |  |  |  |  |
| Lupelele | ICT+ | Wb123+ | Bm14+ | Bm33+ |
|  | ICT+ | Wb123+ | Bm14+ | Bm33+ |
| **TAS-2** |  |  |  |  |
| Lupelele | ICT+ | Wb123+ | Bm14+ | Bm33+ |
| **TAS-3** |  |  |  |  |
| Coleman (Pago) | FTS+ | Wb123− | Bm14− | Bm33+ |
|  | FTS+ | Wb123+ | Bm14+ | Bm33+ |
|  | FTS+ | Wb123+ | Bm14− | Bm33+ |
|  | FTS+ | Wb123+ | Bm14+ | Bm33+ |
| Alataua-Lua | FTS+ | Wb123+ | Bm14+ | Bm33+ |
|  | FTS+ | Wb123+ | Bm14+ | Bm33+ |
| Lupelele | FTS+ | Wb123+ | Bm14+ | Bm33+ |
| Manulele | FTS+ | Wb123− | Bm14− | Bm33− |
| Matafao | FTS+ | Wb123− | Bm14− | Bm33+ |

ICT= Binax NOW® Filariasis Immunochromatographic test
FTS= Alere^TM^ Filariasis Test Strip
